# Utilising the Palliative Prognostic Index in a mixed non-malignant and malignant patient group to determine prognosis. A general medicine tool for prognostication

**DOI:** 10.1101/2024.07.24.24310939

**Authors:** Joseph Hawkins, Megan Lester

**Author notes:** Contributor 1: Susan Dargan, Nurse Consultant, Ashford and St Peter’s NHS Foundation Trust, Nurse lead for Palliative Care. Competing Interests: none declared.

## Abstract

**Objectives:** This study tested the use of the Palliative Prognostic Index (PPI), an established cancer prognostic tool, in a general medicine group within an acute setting for non-selective adult palliative care. The PPI score ranges from 0 to 15, with scores <6 indicating a prognosis of over 6 weeks and scores >6 indicating under 3 weeks.

**Methods:** Data from 256 patients seen over three months by the Ashford and St Peter’s NHS Foundation Trust Palliative Care team were analysed. PPI scores were calculated and correlated with patient’s date of death (DoD) to evaluate predictive value. ASPH is a medium sized hospital in England with 500 adult beds.

**Results:** Among 256 patients, 145 had cancer and 111 had non-malignant disease. Higher PPI scores correlated with more accurate prognostic predictions, with an overall prediction accuracy of 70%.

**Conclusions:** The study demonstrates the PPI tool’s value for mixed groups of non-malignant and malignant diseases. The ASPH population is representative of most UK areas, suggesting that the PPI tool can guide timely care decisions in general medical settings.

## Objectives

Historically, death has been a social phenomenon, with grieving recognised as a part of life. People have always known they would die, and witnessing death has been crucial for societal coping. However, there is an increasing medicalization of dying, and despite patient wishes^1^, hospital remains the most common place of death with approximately 43% of the population dying in hospital^2^. Recognition of dying has become a medical task in today’s health system. The national picture shows pressure on healthcare providers from rising patient expectations and reducing finical flexibility.

The Palliative Prognostic Index (PPI) was developed in 1999 by Morita et al, to support prognostication in cancer patients. The PPI score is calculated by summing scores of five independently predictive variables: PPS scores, oral intake, oedema, dysponea at rest and delirium. PPS scores in this study were transferred from KPS scores, with KPS scores of 10-100 corresponding to PPS scores of 10-100. PPI scores range from 0-15.

## Methods

Data was gathered from every patient seen in a three-month period (256) by the Ashford and St Peter’s NHS Foundation Trust Palliative Care Team. A PPI value was calculated for each patient between 18.5.2021 and 13.8.2021, and then correlated with their date of death (DoD). ASPH is a medium-sized hospital in England with 500 adult beds, Data analysis used StatPlus software for p-values, graphical representations, and predictive value.

## Results

Of 256 patients, 145 were identified as dying of cancer and 111 with non-malignant disease. Among non-malignant diseases, dementia was most common (315), followed by stroke (16%) and heart failure (14%). PPI scores greater then 6 were found in 42% (107) of patients, while 58% (149) had scores fewer then 6. Higher PPI results correlated with increased accuracy of prognostic prediction (P-0.0705) – see graph 1. The highest predictive correlation with DoD was in those with a score of 15 (accurate in 82% of cases), while the lowest was in those with a score of 2 or fewer (accurate in 62% of cases). Overall accuracy was 70% across both malignant and non-malignant groups.

## Conclusions

The study demonstrates the value of PPI tool in general medical population of adult patients with both non-malignant and malignant diseases. The ASPH population reflects typical disease variation, gender and age in the UK. The PPI tool can assist hospitals and hospices by providing an objective prognostication system applicable to as broad population. It supports resource allocation, appropriate place of care, and end of life planning, ensuring timely decisions. Further research is needed on long-term prognostication tools to aid clinical teams in decision-making and adjust therapies for an increasingly frail and comorbid population.

**Figure 1.**
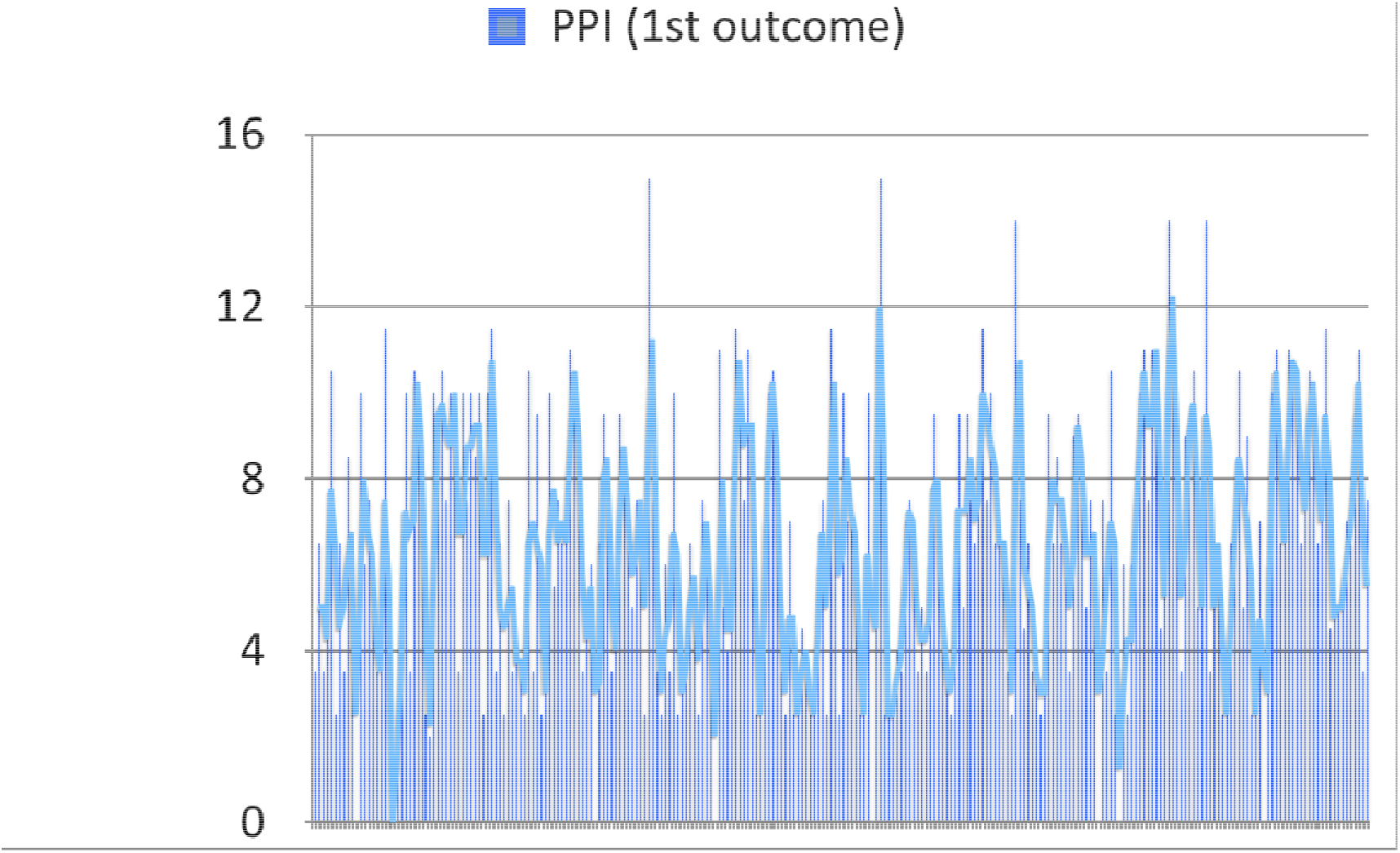
Visual representation of total PPI variability over 3 month period

## Data Availability

All data produced in the present study are available upon reasonable request to the authors

## Notes

### Competing Interest Statement

The authors have declared no competing interest.

### Funding Statement

This study did not receive any funding

### Author Declarations

Ethical Comitte Ashford and St Peter's Foundation Trust

## References

1. Source-national VOICES data

2. https://www.gov.uk/government/statistics/palliative-and-end-of-life-care-profiles-december-2023-data-update/palliative-and-end-of-life-care-profile-december-2023-update-statistical-commentary

